# A Framework for SARS-CoV-2 Testing on a Large University Campus: Statistical Considerations

**DOI:** 10.1101/2020.07.23.20160788

**Authors:** Paul J. Rathouz, Catherine A. Calder

## Abstract

We consider testing strategies for active SARS-CoV-2 infection for a large university community population, which we define. Components of such a strategy include *individuals* tested because they self-select or are recommended for testing by a health care provider for their own health care; individuals tested because they belong to a *high-risk group* where testing serves to disrupt transmission; and, finally, individuals randomly selected for testing from the university community population as part of a *proactive community testing*, or surveillance, program. The proactive community testing program is predicated on a mobile device application that asks individuals to self-monitor COVID-like symptoms daily. The goals of this report are (i) to provide a framework for estimating prevalence of SARS-CoV-2 infection in the university community wherein proactive community testing is a major component of the overall strategy, (ii) to address the issue of how many tests should be performed as part of the proactive community testing program, and (iii) to consider how effective proactive community testing will be for purposes of detection of new disease clusters.

We argue that a comprehensive prevalence estimate informed by *all* testing done of the university community is a good metric to obtain a global picture of campus SARS-CoV-2 infection rates at a particular point in time and to monitor the dynamics of infection over time, for example, estimating the population-level reproductive number, *R*_0_). Importantly, the prevalence metric can be useful to campus leadership for decision making. One example involves comparing campus prevalence to that in the broader off-campus community. We also show that under some reasonable assumptions, we can obtain valid statements about the comprehensive prevalence by only testing symptomatic persons in the proactive community testing component.

The number of tests performed for *individual-level* and *high-risk group-level* needs will depend on the disease dynamics, individual needs, and testing availability. For purposes of this report, we assume that, for these groups of individuals, inferential precision — that is, the accuracy with which we can estimate the true prevalence from testing a random sample of individuals — does *not* drive decisions on the number of tests.

On the other hand, for proactive community testing, the desired level of inferential precision in a fixed period of time can be used to justify the number of tests to perform in that period. For example, our results show that, if we establish a goal of ruling out with 98% confidence a background prevalence of 2% in a given week, and the actual prevalence is 1% among those eligible for proactive community testing, we would need to test 835 randomly-selected symptomatics (i.e., those presenting with COVID-like symptoms) per week via the proactive community testing program in a campus of 80k individuals. In addition to justifying decisions about the number of tests to perform, inferential precision can formalize the intuition that testing of symptomatic individuals should be prioritized over testing asymptomatic individuals in the proactive community testing program.

## 1 Introduction: A Framework for Campus Surveillance

We consider a university population of students, staff, and faculty. We assume the institution has an approximately complete census of persons who are allowed to be on campus. Excluded are members of the community who never come to campus, i.e., working or learning completely remotely. We consider a given point, or narrow window, in time, e.g., a week, and assume that the window is narrow enough that the epidemic is in approximately steady state during that window.

For purposes of this report, we assume a comprehensive campus testing strategy with **three broad objectives** and corresponding purposes of testing:

1. Individuals are tested as a part of evaluation and management of COVID-19. This objective is fully focused at the level of individual health care.
2. Group-level testing is done as a part of contact tracing or cluster delineation to disrupt transmission from an identified source or event. This objective is focused on containment of identified outbreaks.
3. Testing is done to monitor the global — or comprehensive — prevalence of SARS-CoV-2 infection within the population in order to guide population-level decisions aimed at reducing transmission.

These objectives are ordered based on the focus, from the individual to the population. The testing done in support of Objectives 1 and 2 is likely not sufficient for the population inference in Objective 3 (i.e., estimating the prevalence in a large population). Nevertheless, results of testing done in support of 1 and 2, combined with testing done in support of 3, can give us an understanding of prevalence, especially with respect to change over time.

In addition to elaborating a framework for testing or estimating prevalence, this report aims to **quantify via statistical power and sample size calculations** how effective both asymptomatic and symptomatic testing are at monitoring SARS-CoV-2 infection prevalence; **namely at achieving Objective 3**. We consider Objectives 1 and 2 as they impact on global prevalence monitoring. Regarding Objective 2, we also examine how effective symptomatic testing is at detection of clusters of SARS-CoV-2 infection, by quantifying how large a cluster must be until is is expected to be detected with some high probability.

Our approach is first to show how the **three objectives of a testing strategy implicitly define subgroups** of the campus community for each of which we can obtain estimates of SARS-CoV-2 prevalence. The subgroup estimates of prevalence can be considered separately, or we can combine them via weighting based on relative population sizes to determine an overall estimated prevalence that can be monitored over time for use in decision making. For subgroups for which the decision to test is based on random selection of individuals, we consider a formal inferential decision problem to answer the question “How many people need to be tested?” to be able to have sufficient information on which to base decisions.

Note: This report is designed to be consumed without referring to the technical material in the **Statistical Framework and Methods** Section.

## 2 Components of a comprehensive testing strategy

We assume there is a mobile device application (mobile app) for symptom tracking. Everyone who goes to campus is supposed to use the mobile app daily to attest to whether they are experiencing COVID-like symptoms. Individuals who report COVID-like symptoms on the mobile app will be required to stay home or in isolation. In addition, all or a high fraction of those with symptoms meeting a pre-specified set of criteria (symptomatic) will be notified that they should present to be tested. A smaller fraction of those with lesser and/or no symptoms (asymptomatic) may be randomized to receive a test as part of the proactive community testing program.

Based on the testing strategy and prior results, the known campus population in any given week can be divided into subgroups as follows:

**Group A** Individuals who have already been identified in prior weeks as having active SARS-CoV-2 infections, whether symptomatic or not.

**Group B** Individuals who are tested or who report results of tests obtained on their own. These individuals may present for testing on their own, e.g., to the university health service generally used by students, or may be tested based on the advice of a health care professional or for screening purposes prior to hospital admission.

**Group C** Individuals who are tested because they were notified that they were in contact with someone who is SARS-CoV-2 positive, e.g., through contact tracing, or who otherwise belong to a well-defined group of high-risk individuals (e.g., athletes).

**Group D** Individuals who complete the symptom tracker on the mobile app and are eligible to be randomly selected for testing, i.e., who do not fall in any of **Groups A, B**, or **C**. This group comprises two subgroups: **Subgroup D**.**s:** The subgroup of **D** who have COVID-like symptoms. **Subgroup D**.**a:** The subgroup of **D** who do not have COVID-like symptoms (are asymptomatic).

Note that the groups are **highly aligned** with the **three broad objectives** of a testing strategy; in particular, **Groups B, C**, and **D** support Objectives 1, 2, and 3 respectively. In addition, there are several critical features of this framework that allow us to make progress toward comprehensive prevalence monitoring:

- First, each group has a known denominator; that is, we know how many individuals are in each group during each week. This is not necessarily true for the subgroups **D**.**s** and **D**.**a**, but it is true of the main Group **D**. We will handle this problem below.
- A second feature is that the SARS-CoV-2 infection prevalence will be known in each of groups **A**-**C**, but not in **D**. For **A**, the prevalence is 1.0 because these are the previously identified positives. For **B** and **C**, by definition of the group, **all members** will be tested, so group-specific prevalence can be computed directly up to test sensitivity.
- Third, the group specific prevalences can be combined into a global prevalence by taking an average across the groups weighted by their group size.
- **Group D** provides an estimate of the **background prevalence** of SARS-CoV-2 prevalence. This forms part of an estimate of the global prevalence.
- In addition, the **approach can be extended or modified** by **changing the definition of the groups** according to needs specified by university leadership. For example other high risk groups (e.g., an athletic team or a specific dormitory) may be added, and these can be changed week-to-week as needed.

A proactive community testing program can include sampling symptomatics or asymptomatics, or a combination of the two:

### Symptomatic testing

This involves randomly sampling and testing individuals for SARS-CoV-2 infection among those in the population with COVID-like symptoms as identified through the mobile app, i.e., **Group D**.**s**. Under some assumptions, the results from symptomatic testing can be used to *infer* the prevalence of *asymptomatic* SARS-CoV-2 infection as well; we show how later in this report. One such assumption is on the ratio of asymptomatic to symptomatic persons with SARS-CoV-2 infections. Generally, the more comprehensive the program of symptomatic testing, i.e., the larger and the more representative the sample size, the more informative it will be for making inferences about the prevalence of SARS-CoV-2 infection in **Group D**.

### Asymptomatic testing

This involves randomly sampling and testing individuals for SARS-CoV-2 infection among those in the population without COVID-like symptoms, i.e., **Group D**.**a**. Such a program requires a formal system of sampling or selecting individuals for testing, performing symptom surveillance on them (to ensure they do not belong in the symptomatic population, **Group D**.**s**), and recruiting them to present for testing. Exploiting an assumption about the ratio of the number of asymptomatic to number of symptomatic SARS-CoV-2 infections, results can be used to *infer* the prevalence of SARS-CoV-2 infection **Group D**; we show how later in this report. We find that asymptomatic testing is inefficient from a testing volume perspective.

Focusing on these two components specifically, in the next section of this report, we show how and quantify the degree to which symptomatic and asymptomatic testing (**Groups D**.**s** and **D**.**a**) can each be used to make inferences via statistical hypothesis testing about the background prevalence of SARS-CoV-2 infection in the university community. Besides laying out the statistical principles, a main goal here is to **estimate the number of tests** needed for robust inferences on the background prevalence, for **purposes of design of the overall strategy** and **planning for testing capacity**. Of course, as stated, this background prevalence can be combined with results from the other groups to obtain estimates of the comprehensive — or global —prevalence.

## 3 Proactive community testing

### 3.1 A statistical framework

Focusing here on background prevalence testing in **Group D** alone, the statistical framework in the **Methods** highlights the challenges in *estimation* of prevalence of SARS-CoV-2 infection in an unbiased way. To do so will either require representative data from both the symptomatic and the asymptomatic populations and/or accurate assumptions about parameters such as the ratio of symptomatic to asymptomatic persons with SARS-CoV-2 infections and/or the current prevalence of COVID-like symptoms.

As such, rather than pursuing estimation, we show how to *statistically test* for an *upper bound on prevalence*, even if we cannot obtain an accurate estimate thereof. The idea behind the approach, when applied, is for university health and safety leaders to specify as a threshold the *highest* SARS-CoV-2 prevalence that is *acceptably safe*, and to design a strategy that will rule out prevalence at that level (or higher) with high probability when the prevalence is in fact lower than the threshold. Here, as a starting point, we consider asymptomatic and symptomatic testing separately. For each, we aim to identify *how many tests* are needed and what the *critical number of positives* will be, below which we rule out prevalence above the threshold.

#### Asymptomatic testing

Consider a true but unknown overall prevalence *π*, and threshold-of-safety prevalence *π*^0^. We show in the **Methods** that we can test *π* = *π*^0^ versus *π < π*^0^ by performing SARS-CoV-2 testing on a random sample of *asymptomatics only* (i.e., **Group D**.**a**). To accomplish this, we need for *several design specifications* and *assumptions* to be made by university health and safety officials, as follows. We provide example values here.

- Assumption: The asymptomatic persons selected and presenting for testing are a random sample—or an approximation thereof—of all asymptomatic persons in the population.
- Assumption: Among all undetected SARS-CoV-2 infections, the proportion who are symptomatic is an assumed known quantity. Call this *λ*_*s*_. Here, we assume that *λ*_*s*_ is between 0.25 and 0.40, supported by recent data (Poletti *et al*.; Gostic *et al*.). Note that higher values of *λ*_*s*_ will lead to more conservative strategies in the sense that we will tend to test more than strictly needed. As such, we set *λ*_*s*_ = 0.40.
- The smallest unacceptable prevalence *π*^0^ to rule out. Here, we consider 0.5%, 1%, 2%, 4%, and 8%. These levels may seem high relative to current population estimates; we note, however, that those estimates are often based only on testing in the general population, and it is generally recognized that such testing substantially underestimates the prevalence of active SARS-CoV-2 infections.
- Our best guess of the actual prevalence in the community, *π*^*A*^. Here, we set this value to be one half of the smallest unacceptable prevalence.
- The test sensitivity, *γ*_se_ (i.e., the probability of a positive test result given that the individual being tested is in fact SARS-CoV-2 positive). Here, we set this test characteristic at either 0.70, 0.85.
- The confidence level around *π* = *π*^0^. This is one minus the likelihood of falsely claiming that *π < π*^0^ when it is not true^1^. Owing to the health and safety implications of falsely detecting a low prevalence, we consider fairly high confidence levels of 0.98 and 0.95.
- The target probability of ruling out *π* = *π*^0^ when in fact *π < π*^0^^2^. We would like this probability to be high, but also recognize that it may be constrained by available resources. Here, we consider a value of 90%.

We show in the **Methods** that testing in **Group D** as a whole can be accomplished via testing a random sample of *asymptomatics only* and ruling out via hypothesis testing 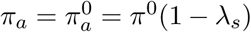, or anything greater.

#### Symptomatic testing

Alternatively, we could perform testing on a random sample of *symptomatics only*—persons with COVID-like symptoms. As above, we need for *several design specifications* and *assumptions* to be made by university health and safety officials, as follows.

- Assumption: The persons selected and presenting for testing are a random sample—or an approximation thereof—of the symptomatic persons in the population.
- Assumption: Again, among all undetected SARS-CoV-2 infections, the proportion who are symptomatic is an assumed known quantity *λ*_*s*_. We assumed above that *λ*_*s*_ is between 0.25 and 0.40; however, with symptomatic testing, lower values lead to more required tests, so we set *λ*_*s*_ = 0.25 to be sure to have enough to draw valid conclusions.
- Assumption: The prevalence in the population of persons with COVID-like symptoms, *ν*_*s*_, is an assumed known quantity. Higher values will lead to more tests. Here, we assume these values range between 2% and 9%, using crude application of ranges in the following, and set *ν*_*s*_ = 9%.

- Symptoms due to rhinovirus: Estimates ranged from 2% to 4% based on Eggo *et al*.
- Symptoms due to influenza: In the 2018-19 flu season, CDC reported that about 35 million people had flu (about 10% of the U.S. population) over a period of about 5 months (October to February). Assuming symptoms may last up to 2 weeks, that would be about 1% at any given time. So, symptom prevalence due to flu would range from 0% to 1%.
- Symptoms due to COVID-19: The low end would be 0.5% prevalence times 25% of infected being symptomatic, yielding 0.125%. The high end would be 8% prevalence with 40% symptomatic, yielding 3.2%.

With such assumptions, we show in the **Methods** that testing for prevalence can be accomplished via testing a random sample of *symptomatics only* and ruling out via hypothesis testing 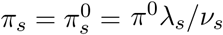, or anything greater.

### 3.2 Application and Results

To obtain a sense of the required volume of testing needed, consider testing in **Group D** alone, either asymptomatic (**D**.**a**) or symptomatic (**D**.**s**).

#### Asymptomatic testing

Consider a program of asymptomatic testing in a randomly selected sample with a minimum sample size of 800/week, justified as follows. The campus community comprises about 100k individuals, 80k of whom will be on campus at some point. Testing 1.0% of the population per week requires 800 tests/week.

Table 1 contains the critical values (the largest number of positive tests under which we would still rule out *π* = *π*^0^) and power (the probability of ruling out *π* = *π*^0^ when in fact *π < π*^0^) for 800 tests/week. To interpret Table 1, consider Row 13. If our threshold for safety is 4% prevalence of SARS-CoV-2 infection, then we would rule that level out with 98% confidence if we observe 6 or fewer positives out of a sample of 800. However, if the true prevalence is in fact 2%, then our chances of seeing 6 or fewer positives is only 49%. Indeed, the NA values in Table 1 indicate that the desired confidence of 95% or 98% cannot be obtained at that sample size. Overall, at this testing level, the power is unacceptably low for all but the highest prevalence level of 8%.

**Table 1:** One-sided critical values and power for *m*_*a*•_ = 800 asymptomatic tests per week.

|  | lambda.s | pi0 | gamma.se | conf | piA | crit | power |
| --- | --- | --- | --- | --- | --- | --- | --- |
| 1 | 0.4 | 0.005 | 0.70 | 0.98 | 0.0025 | NA | NA |
| 2 | 0.4 | 0.005 | 0.70 | 0.95 | 0.0025 | NA | NA |
| 3 | 0.4 | 0.005 | 0.85 | 0.98 | 0.0025 | NA | NA |
| 4 | 0.4 | 0.005 | 0.85 | 0.95 | 0.0025 | NA | NA |
| 5 | 0.4 | 0.010 | 0.70 | 0.98 | 0.0050 | NA | NA |
| 6 | 0.4 | 0.010 | 0.70 | 0.95 | 0.0050 | 0 | 0.19 |
| 7 | 0.4 | 0.010 | 0.85 | 0.98 | 0.0050 | 0 | 0.13 |
| 8 | 0.4 | 0.010 | 0.85 | 0.95 | 0.0050 | 0 | 0.13 |
| 9 | 0.4 | 0.020 | 0.70 | 0.98 | 0.0100 | 1 | 0.15 |
| 10 | 0.4 | 0.020 | 0.70 | 0.95 | 0.0100 | 2 | 0.35 |
| 11 | 0.4 | 0.020 | 0.85 | 0.98 | 0.0100 | 2 | 0.23 |
| 12 | 0.4 | 0.020 | 0.85 | 0.95 | 0.0100 | 3 | 0.42 |
| 13 | 0.4 | 0.040 | 0.70 | 0.98 | 0.0200 | 6 | 0.49 |
| 14 | 0.4 | 0.040 | 0.70 | 0.95 | 0.0200 | 7 | 0.64 |
| 15 | 0.4 | 0.040 | 0.85 | 0.98 | 0.0200 | 8 | 0.57 |
| 16 | 0.4 | 0.040 | 0.85 | 0.95 | 0.0200 | 9 | 0.70 |
| 17 | 0.4 | 0.080 | 0.70 | 0.98 | 0.0400 | 16 | 0.80 |
| 18 | 0.4 | 0.080 | 0.70 | 0.95 | 0.0400 | 18 | 0.91 |
| 19 | 0.4 | 0.080 | 0.85 | 0.98 | 0.0400 | 21 | 0.90 |
| 20 | 0.4 | 0.080 | 0.85 | 0.95 | 0.0400 | 23 | 0.96 |

lambda.s: proportion in the population who are symptomatic.
pi0: threshold-of-safety prevalence of SARS-CoV-2 infection.
gamma.se: sensitivity of SARS-CoV-2 test.
conf: confidence level for ruling out prevalence equal to or greater than pi0.
piA: actual prevalence.
crit: critical value: number of positives at or below which pi0 can be ruled out.
power: probability of strategy ruling out pi0 when true value is piA

Table 2 estimates the sample size needed (above 800) to attain the desired power of 90%. It also presents critical values and actual power^3^. Consider again Row 13. This says that if we do 2012 tests and obtain 22 or fewer positives, then we rule out prevalence of 4% or higher with 98% confidence. If our actual prevalence is 2%, we have 91% power to see 22 or fewer positives. In Table 2, the NA values indicate that the target power was not attainable with sample sizes less than 4000. Except for the highest levels of prevalence, sample sizes needed to achieve the specified testing targets are quite high.

**Table 2:** Sample sizes *m*_*a*•_ per week, critical values, and realized power with target of 80% for one-sided asymptomatic tests.

|  | lambda.s | pi0 | gamma.se | conf | piA | m.aDot | crit | power |
| --- | --- | --- | --- | --- | --- | --- | --- | --- |
| 1 | 0.4 | 0.005 | 0.70 | 0.98 | 0.0025 | NA | NA | NA |
| 2 | 0.4 | 0.005 | 0.70 | 0.95 | 0.0025 | NA | NA | NA |
| 3 | 0.4 | 0.005 | 0.85 | 0.98 | 0.0025 | NA | NA | NA |
| 4 | 0.4 | 0.005 | 0.85 | 0.95 | 0.0025 | NA | NA | NA |
| 5 | 0.4 | 0.010 | 0.70 | 0.98 | 0.0050 | NA | NA | NA |
| 6 | 0.4 | 0.010 | 0.70 | 0.95 | 0.0050 | NA | NA | NA |
| 7 | 0.4 | 0.010 | 0.85 | 0.98 | 0.0050 | NA | NA | NA |
| 8 | 0.4 | 0.010 | 0.85 | 0.95 | 0.0050 | NA | NA | NA |
| 9 | 0.4 | 0.020 | 0.70 | 0.98 | 0.0100 | NA | NA | NA |
| 10 | 0.4 | 0.020 | 0.70 | 0.95 | 0.0100 | 3032 | 17 | 0.90 |
| 11 | 0.4 | 0.020 | 0.85 | 0.98 | 0.0100 | 3317 | 22 | 0.91 |
| 12 | 0.4 | 0.020 | 0.85 | 0.95 | 0.0100 | 2496 | 17 | 0.91 |
| 13 | 0.4 | 0.040 | 0.70 | 0.98 | 0.0200 | 2012 | 22 | 0.91 |
| 14 | 0.4 | 0.040 | 0.70 | 0.95 | 0.0200 | 1514 | 17 | 0.91 |
| 15 | 0.4 | 0.040 | 0.85 | 0.98 | 0.0200 | 1656 | 22 | 0.91 |
| 16 | 0.4 | 0.040 | 0.85 | 0.95 | 0.0200 | 1246 | 17 | 0.91 |
| 17 | 0.4 | 0.080 | 0.70 | 0.98 | 0.0400 | 967 | 21 | 0.90 |
| 18 | 0.4 | 0.080 | 0.70 | 0.95 | 0.0400 | 800 | 18 | 0.91 |
| 19 | 0.4 | 0.080 | 0.85 | 0.98 | 0.0400 | 825 | 22 | 0.91 |
| 20 | 0.4 | 0.080 | 0.85 | 0.95 | 0.0400 | 800 | 23 | 0.96 |

m.aDot: minimum number of tests needed to meet test specifications. see Table 1 for other notes.

#### Symptomatic testing

Consider a program of symptomatic testing involving a representative sample of 3200 tests per week. Whereas we view this as a maximum number, it is not unrealistic under our assumptions about the prevalence of of COVID-like symptoms being between 2% and 9%. If that level is 4%, and there are 80k persons in the population, and all symptomatics present for testing, then 3200 tests will be required.

Analogous to Table 1, Table 3 contains the critical values and power for 3200 tests/week on a random sample of symptomatics. The power is quite high. Table 4 presents sample sizes up to 3200, and associated critical values, to achieve at least 90% power. Sample sizes are modest for most cases, and are considerably less than under an asymptomatic testing program. For example, see line 9: To rule out with *>*98% confidence a background prevalence of 2% when the actual prevalence is 1%, we require 835 tests per week among randomly-selected symptomatics via the proactive community testing program.

**Table 3:** One-sided critical values and power for *m*_*s*•_ = 3200 symptomatic tests per week. see Table 1 for notes.

|  | lambda.s | nu.s | pi0 | gamma.se | conf | piA | crit | power |
| --- | --- | --- | --- | --- | --- | --- | --- | --- |
| 1 | 0.25 | 0.09 | 0.005 | 0.70 | 0.98 | 0.0025 | 19 | 0.84 |
| 2 | 0.25 | 0.09 | 0.005 | 0.70 | 0.95 | 0.0025 | 21 | 0.93 |
| 3 | 0.25 | 0.09 | 0.005 | 0.85 | 0.98 | 0.0025 | 25 | 0.93 |
| 4 | 0.25 | 0.09 | 0.005 | 0.85 | 0.95 | 0.0025 | 27 | 0.97 |
| 5 | 0.25 | 0.09 | 0.010 | 0.70 | 0.98 | 0.0050 | 46 | 1.00 |
| 6 | 0.25 | 0.09 | 0.010 | 0.70 | 0.95 | 0.0050 | 49 | 1.00 |
| 7 | 0.25 | 0.09 | 0.010 | 0.85 | 0.98 | 0.0050 | 57 | 1.00 |
| 8 | 0.25 | 0.09 | 0.010 | 0.85 | 0.95 | 0.0050 | 61 | 1.00 |
| 9 | 0.25 | 0.09 | 0.020 | 0.70 | 0.98 | 0.0100 | 101 | 1.00 |
| 10 | 0.25 | 0.09 | 0.020 | 0.70 | 0.95 | 0.0100 | 106 | 1.00 |
| 11 | 0.25 | 0.09 | 0.020 | 0.85 | 0.98 | 0.0100 | 126 | 1.00 |
| 12 | 0.25 | 0.09 | 0.020 | 0.85 | 0.95 | 0.0100 | 131 | 1.00 |
| 13 | 0.25 | 0.09 | 0.040 | 0.70 | 0.98 | 0.0200 | 217 | 1.00 |
| 14 | 0.25 | 0.09 | 0.040 | 0.70 | 0.95 | 0.0200 | 223 | 1.00 |
| 15 | 0.25 | 0.09 | 0.040 | 0.85 | 0.98 | 0.0200 | 268 | 1.00 |
| 16 | 0.25 | 0.09 | 0.040 | 0.85 | 0.95 | 0.0200 | 274 | 1.00 |
| 17 | 0.25 | 0.09 | 0.080 | 0.70 | 0.98 | 0.0400 | 455 | 1.00 |
| 18 | 0.25 | 0.09 | 0.080 | 0.70 | 0.95 | 0.0400 | 463 | 1.00 |
| 19 | 0.25 | 0.09 | 0.080 | 0.85 | 0.98 | 0.0400 | 558 | 1.00 |
| 20 | 0.25 | 0.09 | 0.080 | 0.85 | 0.95 | 0.0400 | 567 | 1.00 |

**Table 4:** Sample sizes *m*_*a*_ per week, critical values, and realized power with target of 90% for one-sided symptomatic tests.

|  | lambda.s | nu.s | pi0 | gamma.se | conf | piA | m.sDot | crit | power |
| --- | --- | --- | --- | --- | --- | --- | --- | --- | --- |
| 1 | 0.25 | 0.09 | 0.005 | 0.70 | 0.98 | 0.0025 | NA | NA | NA |
| 2 | 0.25 | 0.09 | 0.005 | 0.70 | 0.95 | 0.0025 | 2619 | 17 | 0.91 |
| 3 | 0.25 | 0.09 | 0.005 | 0.85 | 0.98 | 0.0025 | 2865 | 22 | 0.91 |
| 4 | 0.25 | 0.09 | 0.005 | 0.85 | 0.95 | 0.0025 | 2156 | 17 | 0.91 |
| 5 | 0.25 | 0.09 | 0.010 | 0.70 | 0.98 | 0.0050 | 1737 | 22 | 0.91 |
| 6 | 0.25 | 0.09 | 0.010 | 0.70 | 0.95 | 0.0050 | 1308 | 17 | 0.91 |
| 7 | 0.25 | 0.09 | 0.010 | 0.85 | 0.98 | 0.0050 | 1378 | 21 | 0.90 |
| 8 | 0.25 | 0.09 | 0.010 | 0.85 | 0.95 | 0.0050 | 1076 | 17 | 0.91 |
| 9 | 0.25 | 0.09 | 0.020 | 0.70 | 0.98 | 0.0100 | 835 | 21 | 0.90 |
| 10 | 0.25 | 0.09 | 0.020 | 0.70 | 0.95 | 0.0100 | 652 | 17 | 0.91 |
| 11 | 0.25 | 0.09 | 0.020 | 0.85 | 0.98 | 0.0100 | 686 | 21 | 0.90 |
| 12 | 0.25 | 0.09 | 0.020 | 0.85 | 0.95 | 0.0100 | 536 | 17 | 0.91 |
| 13 | 0.25 | 0.09 | 0.040 | 0.70 | 0.98 | 0.0200 | 415 | 21 | 0.91 |
| 14 | 0.25 | 0.09 | 0.040 | 0.70 | 0.95 | 0.0200 | 309 | 16 | 0.90 |
| 15 | 0.25 | 0.09 | 0.040 | 0.85 | 0.98 | 0.0200 | 328 | 20 | 0.90 |
| 16 | 0.25 | 0.09 | 0.040 | 0.85 | 0.95 | 0.0200 | 254 | 16 | 0.90 |
| 17 | 0.25 | 0.09 | 0.080 | 0.70 | 0.98 | 0.0400 | 189 | 19 | 0.90 |
| 18 | 0.25 | 0.09 | 0.080 | 0.70 | 0.95 | 0.0400 | 145 | 15 | 0.90 |
| 19 | 0.25 | 0.09 | 0.080 | 0.85 | 0.98 | 0.0400 | 155 | 19 | 0.91 |
| 20 | 0.25 | 0.09 | 0.080 | 0.85 | 0.95 | 0.0400 | 119 | 15 | 0.90 |

m.aDot: minimum number of tests needed to meet test specifications. see Table 1 for other notes.

### 3.3 Example: Testing week by week

Given this framework, to see how this might work in practice, consider the strategy prescribed in line 9 of Table 4 for the proactive community testing program. Recall that if we establish a goal of ruling out with 98% confidence a background prevalence of 2%, and the actual prevalence is 1% among those in Group D, we would need to test 835 per week randomly-selected symptomatics (i.e., those presenting with COVID-like symptoms) via the proactive community testing program.

Suppose that over six (6) weeks, we observe the following counts of positives among the 835 tested:

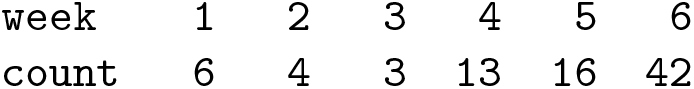

|  |  |  |  |  |  |  |
| --- | --- | --- | --- | --- | --- | --- |
| **week** | **1** | **2** | **3** | **4** | **5** | **6** |
| **count** | 6 | 4 | 3 | 13 | 16 | 42 |

How would these data be used to drive decisions? As an example, assume the values *λ*_*s*_ = 0.25, *ν*_*s*_ = 0.09, *γ*_se_ = 0.70. With these parameters and “data,” Figure 1 shows the estimated prevalence among the symptomatics (open blue triangles, corrected for imperfect test sensitivity), the derived estimated global prevalence (closed blue triangles), and, critically, the upper 98% confidence bound for the global prevalence (closed red triangles).

**Figure 1:**
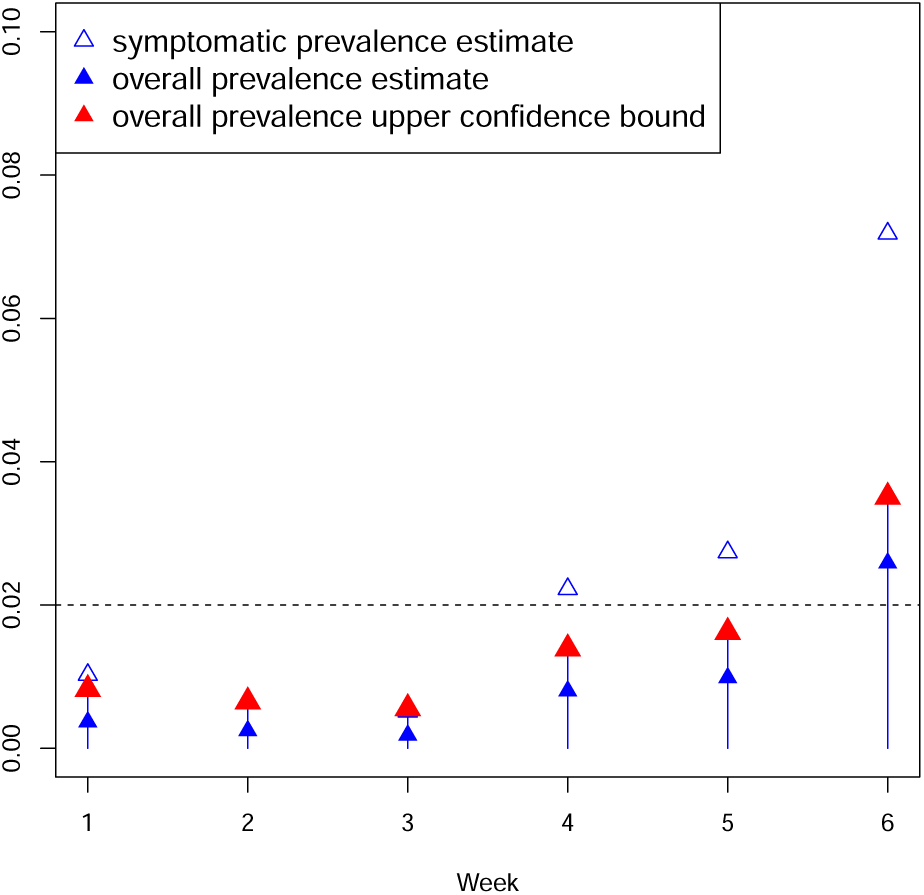
Hypothetical prevalence estimates and upper bound for six weeks of a proactive community testing program. The estimated prevalence (corrected for imperfect test sensitivity) among the symptomatics is represented by open blue triangles; the derived estimated global prevalence in Group **D** is represented by closed blue triangles along with 98% one-sided confidence intervals represented by blue vertical lines; and the upper 98% confidence bound for the global prevalence is presented by closed red triangles.

Recall that for 98% confidence bounds, we would need to see 21 positives or less to rule out a global prevalence of 2% or more. We see that the confidence bound is below 2% for the count of 16 — indeed in this example, the estimated overall prevalence is not much more than 1% until week 6 — but not for the count of 42. This would presumably trigger a decision in Week 6.

Owing to inequalities in assumed parameter values, these are are likely biased high; for the upper bounds in the red triangles, however, this is acceptable because it will lead to decisions that will err on the side of safety.

## 4 Cluster Detection

In the foregoing, we have largely considered testing needed to achieve the third objective of a testing program, namely monitoring the global prevalence of active SARS-CoV-2 infection in the campus community. Whereas this is an important baseline metric to track and control, it is likely insufficient to maintain safety and to proactively contain transmission of infection. As mentioned in the **Introduction**, for this purpose, identification and containment of outbreaks is a critical component of a comprehensive testing program. We might therefore ask how effective sampling and testing among the population with COVID-like symptoms is at cluster detection.

Specifically, sampling of the symptomatic group, namely group **D**.**s**, via *proactive community testing* allows for **cluster detection** in the following way. Consider a given (as yet undetected) cluster within the campus community. For this simple analysis, we ignore the connection of that cluster to any other cluster. We consider a cluster to be *detected* when one member of that cluster tests positive, so that contact tracing can begin. How effective is proactive community testing at detecting the cluster? For planning purposes, we answer that question by quantifying how big the cluster needs to be before it has a high probability of being detected. This is similar to an approach taken by Martin *et al*., although those authors approach the problem via simulations in the context of a mathematical epidemiologic spread model.

When randomly testing within population **D**.**s**, a SARS-CoV-2 positive individual is detected if s/he has symptoms (with probability *λ*_*s*_, the proportion of SARS-COV-2 infected persons who are symptomatic), if s/he is randomly selected from **Group D**.**s** for testing given s/he has symptoms (with probability equal to the number of tests conducted divided by number with COVID-like symptoms in the population), and if s/he tests positive given she is positive (with test sensitivity *γ*_se_). Given the size of the cluster, that cluster is *not* detected if none of members are detected, and we can use this to compute the probability of the cluster being detected. In Table 5, assuming a population of 3200 with COVID-like symptoms, we consider a variety of scenarios and provide the size to which the cluster must grow before it has a 90% or a 95% chance of being detected in a given week.

**Table 5:** Minium cluster sizes needed under various scenarios to achieve a high probability of detection via proactive community testing of the population with COVID-like symptoms.

|  | m.sDot | lambda.s | gamma.se | det.prob | size |
| --- | --- | --- | --- | --- | --- |
| 1 | 800 | 0.25 | 0.70 | 0.95 | 67 |
| 2 | 800 | 0.25 | 0.70 | 0.90 | 52 |
| 3 | 800 | 0.25 | 0.85 | 0.95 | 55 |
| 4 | 800 | 0.25 | 0.85 | 0.90 | 43 |
| 5 | 800 | 0.40 | 0.70 | 0.95 | 42 |
| 6 | 800 | 0.40 | 0.70 | 0.90 | 32 |
| 7 | 800 | 0.40 | 0.85 | 0.95 | 34 |
| 8 | 800 | 0.40 | 0.85 | 0.90 | 26 |
| 9 | 1600 | 0.25 | 0.70 | 0.95 | 33 |
| 10 | 1600 | 0.25 | 0.70 | 0.90 | 26 |
| 11 | 1600 | 0.25 | 0.85 | 0.95 | 27 |
| 12 | 1600 | 0.25 | 0.85 | 0.90 | 21 |
| 13 | 1600 | 0.40 | 0.70 | 0.95 | 20 |
| 14 | 1600 | 0.40 | 0.70 | 0.90 | 16 |
| 15 | 1600 | 0.40 | 0.85 | 0.95 | 17 |
| 16 | 1600 | 0.40 | 0.85 | 0.90 | 13 |
| 17 | 2400 | 0.25 | 0.70 | 0.95 | 22 |
| 18 | 2400 | 0.25 | 0.70 | 0.90 | 17 |
| 19 | 2400 | 0.25 | 0.85 | 0.95 | 18 |
| 20 | 2400 | 0.25 | 0.85 | 0.90 | 14 |
| 21 | 2400 | 0.40 | 0.70 | 0.95 | 13 |
| 22 | 2400 | 0.40 | 0.70 | 0.90 | 10 |
| 23 | 2400 | 0.40 | 0.85 | 0.95 | 11 |
| 24 | 2400 | 0.40 | 0.85 | 0.90 | 8 |

note: Assume 3200 persons with COVID-like symptoms eligible for sampling.
lambda.s: proportion in the population who are symptomatic.
gamma.se: sensitivity of SARS-CoV-2 test.
det.prob: target probability of detection of cluster
size: minimum cluster size to achieve target probability

Considering sample sizes of 800 to 2400 (out of 3200) per week of the symptomatic population **D**.**s**, we consider ranges of other parameter values in Table 5. Under the most difficult scenario (*λ*_*s*_ = 0.25, *γ*_se_ = 0.70), line 1 in the Table 5 tells us the cluster could grow to 67 SARS-CoV-2 infected individuals before reaching a 95% chance of being detected if we only test 800/3200 per week among those with COVID-like symptoms. Alternatively, if we aggressively test as many symptomatics as possible, and are able to achieve a 75% rate of testing, then cluster size could be as small as 22 before being detected (line 18 in Table 5); even so, it could very well be that cluster sizes in this range, given exponential transmission, are too dangerously high.

These are over-estimates of detectable cluster sizes because they do do not take account of clusters being detected through an individual presenting for testing as part of **Group B** or through new clusters identified through testing in **Group C**; nevertheless, it does give a sense of what is needed containment of transmission of an ongoing epidemic.

## 5 Discussion and Recommendation

In this report, we have first provided a framework based on different objectives and corresponding subpopulations for testing for active SARS-CoV-2 infection in a large university campus setting. Second, focusing on the specific problem of monitoring global — or comprehensive — prevalence of infection via proactive community testing, we provide sample sizes needed to obtain usable results for decision-making, where testing is done in either the asymptomatic or the symptomatic population. Lastly, although briefly, we provided testing proportions among those with COVID-like symptoms needed to detect clusters with high probability before the cluster grows too large.

Our framework assumes active monitoring of COVID-like symptoms in the population via a mobile app. The framework should be augmented by strong messaging about how and when to self-monitor symptoms, and when to get tested.

Whereas we have separated asymptomatic and symptomatic testing for purposes of estimating the number of tests, there will be options to combine the results for an overall test of the prevalence, in the case where both groups are sampled and tested.

Even so, a **key take-away conclusion** from the sample size estimations is that resources should be placed on developing and implementing a **robust testing strategy among the symptomatics** so long as there is a sufficient number of symptomatics who self-identify through mobile app health attestation, and to substantially downplay or eliminate asymptomatic testing. Testing in the population with COVID-like symptoms will also contribute directly to individual-level care and targeted group interventions (isolation, contact tracing) that serve to disrupt transmission, in addition to estimating overall SARS-CoV-2 prevalence.

Given the added benefits of symptomatic testing (beyond prevalence monitoring), and the greater statistical power, we recommend the following: If *testing resources are constrained*, they should all be devoted to the widest program of symptomatic testing possible, including recruiting symptomatics for testing via public health messaging and symptom surveillance. With *more generous resources* or when insufficient numbers of symptomatics are available for proactive community testing, asymptomatic testing could be added, but should not replace symptomatic testing. Asymptomatic testing is also better targeted to *suspected high risk groups* that can be distinguished from the more general asymptomatic population.

Limitations arise in this framework if sampled individuals do not present for testing, if individuals mis- or under-report their symptoms, e.g., through the mobile app, if persons with symptoms do not present for testing, or if adherence to the mobile app requirement is weak. The sample sizes presented are based on the totals actually tested and those individuals comprising a truly representative sample. As such, there are several opportunities for bias in results here. In addition, we have avoided the issue of defining the criteria for having COVID-like symptoms (i.e., being assigned to **Group D**.**s**) versus being asymptomatic (**Group D**.**a**), and have only assumed that a crisp definition can be established based on data from the mobile app. Finally, we have not yet accounted for less than 100% test sensitivity in the testing needed for **Groups B** and **C**. That can be handled at analysis time.

## 6 Statistical Framework and Methods

### Parameters assumed known (or with a known prior)

These are nuisance parameters in the sense that we are not trying to infer on them, but are needed for valid inferences.

- *N_••_*: The total number of persons in the population (on campus), excluding those who are known positives (who we assume are isolated or hospitalized), at the given point in time.
- *γ*_se_: Sensitivity of the test. Say 70% or 85%.
- *η*_*s*_: Among the population with symptoms (s), the proportion of who come in for testing. This parameter is not important if only asymptomatics are tested.
- *λ*_*s*_: Among all undetected infecteds, the proportion who are showing symptoms. Note: This parameter is less important in strategies involving both asymptomatics and symptomatics.

### Parameters needed for design, but not for inferences

These are parameters that will help to create more efficient and informative sampling designs, but which will not invalidate inferences if we get them wrong.

- *η*_*a*_: Among the population who are asymptomatic (a), the proportion who would agree to be tested / show up for testing if asked.
- *π*: The background prevalence of infection among those not known to be positive, **or** the expected null hypothesis value of prevalence.
- *λ*_*s*_: Same as above. This is only needed for design if we are able to test both asymptomatics and symptomatics.

### Finite population counts and sampled counts

Population:

|  |  | COVID-19+ |  |  |
| --- | --- | --- | --- | --- |
|  |  | 1 | 0 | Tot |
| Any Sx | $s$ | $N_{s1}$ | $N_{s0}$ | $N_{s\bullet}$ |
| | $a$ | $N_{a1}$ | $N_{a0}$ | $N_{a\bullet}$ |
| Tot | | $N_{\bullet 1}$ | $N_{\bullet 0}$ | $N_{\bullet\bullet}$ |

where an *N* indicates population counts, 1/0 indicates positive/negative, and *s*/*a* indicates symptomatic or asymptomatic, including among those who are COVID-19 negative.

Sampling among the asymptomatics:

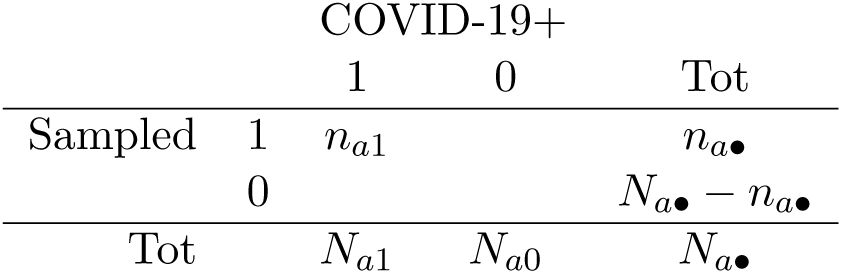

where an *n* indicates sample counts. The **key design parameter** is the number of asymptomatics sampled, viz *n*_*a*•_, but see below where this is replaces by *m*_*a*•_, the number who actually get tested.

### Finite population parameters of interest (targets of interest)

The **main target** is the current prevalence, *N*_•1_*/N*_*•*•_, or equivalently from an inferential perspective, the count *N*_•1_ (because *N*_*•*•_ is assumed known).

It might be useful to target inferences separately on *N*_*s*1_ and *N*_*a*1_.

### Random variables (observed and unobserved)

Observed random variables:

- *m*_*s*•_: The number out of *N*_*s*•_ symptomatics who show up for testing. Assume *for now* that this represents a **random sample** from the *N*_*s*•_ currently active. <u>This is likely a concern</u>.
- 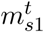: The number out of *m*_*s*1_ positive symptomatics who *test positive* for COVID-19.
- *m*_*a*•_: The number out of *n*_*a*•_ asymptomatics who agree and show up for testing. Assume *for now* that this represents a **random sample** from the *n*_*a*•_ selected. <u>This is likely a concern</u>.
- 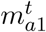: The number out of *m*_*a*1_ positive asymptomatics who *test positive* for COVID-19.

Unobserved random variables (these are unobserved because test sensitivity is not 100%):

- *m*_*s*1_: The number out of *m*_*s*•_ tested symptomatics who are positive for COVID-19.
- *m*_*a*1_: The number out of *m*_*a*•_ tested asymptomatics who are positive for COVID-19.

This gives rise to two new tables

Realized sampling among the symptomatics:

|  |  | COVID-19+ |  |  |
| --- | --- | --- | --- | --- |
|  |  | 1 | 0 | Tot |
| Sampled | 1 | $m_{s1}$ | | $m_{s\bullet}$ |
| | 0 | | | $N_{s\bullet} - m_{s\bullet}$ |
| Tot | | $N_{s1}$ | $N_{s0}$ | $N_{s\bullet}$ |

Realized sampling among the asymptomatics:

|  |  | COVID-19+ |  |  |
| --- | --- | --- | --- | --- |
|  |  | 1 | 0 | Tot |
| Sampled | 1 | $m_{a1}$ | | $m_{a\bullet}$ |
| | 0 | | | $N_{a\bullet} - m_{a\bullet}$ |
| Tot | | $N_{a1}$ | $N_{a0}$ | $N_{a\bullet}$ |

The foregoing is causing me some problems. Without assuming that *all* of those with some symptoms show up for testing, we have no way of counting the number of persons on campus with symptoms, i.e., *N*_*s*•_. That in turn creates challenges to know the value of *N*_*a*•_, the number of persons on campus without symptoms.

Three possible avenues forward here:

- Make assumptions about the ratio *η*_*s*_.
- Make assumptions about the ratio *λ*_*s*_. There is an emerging literature on this.
- Collect data, e.g., via a mobile app, on how many people are showing symptoms.

### Models to be used for inference

These models are based on finite sample inference.

- *m*_*s*•_|*N*_*s*•_ ∼ Bin(*η*_*s*_, *N*_*s*•_)
- *m*_*s*1_|*m*_*s*•_ ∼ HyperG(*N*_*s*•_, *m*_*s*•_, *N*_*s*1_, central)
- 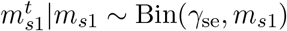
- *m*_*a*1_|*m*_*a*•_ ∼ HyperG(*N*_*a*•_, *m*_*a*•_, *N*_*a*1_, central). This is similar to the second table above, but replaces the number sampled *n*_*a*•_ with the number showing up *m*_*a*•_.
- 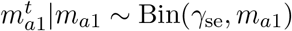

### Statistical simplifications

For purposes of broad outline of the design of testing strategy, and, importantly, determining the testing quantity and frequency for planning purposes, we make several simplifying statistical assumptions and approximations. First, instead of using the statistically correct hypergeometric distribution, we approximate with the binomial distribution. This replaces finite population inference with model-based infinite population inference:

- *m*_*s1*_|*m*_*s*•_ ∼ Bin(*π*_*s*_, *m*_*s*•_),where 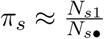
- *m*_*a1*_|*m*_*a*•_ ∼ Bin(*π*_*a*_, *m*_*a*•_),where 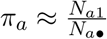

Using the foregoing, we can integrate over *m*_*s*1_ and *m*_*a*1_ to obtain the following, only involving observed quantities:

- 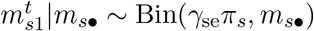
- 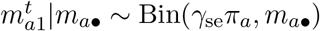

### Asymptomatic only testing

Our immediate aim is to estimate the number of tests needed to rule out given prevalence levels. As one narrow scenario for testing, suppose we just focus on the asymptomatic population, i.e., the population with no COVID-like symptoms. We will leverage assumptions about *λ*_*s*_, the ratio of symptomatic to all cases among infected persons.

Link the prevalence in the asymptomatics to that in the symptomatics:

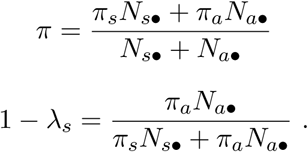

This permits an *upper bound* on *π* as a function of *π*_*a*_:

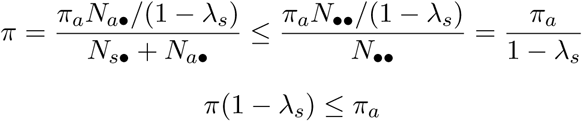

So, for a given null value *π*^0^ of *π*, we can conservatively test (in a statistical hypothesis test) whether *π*_*a*_ is less than a null value of 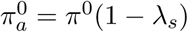. If we reject that hypothesis, then, because

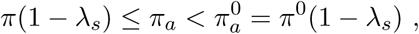

we also reject *π* = *π*^0^ and conclude that *π < π*^0^.

Remarks: (a) To be conservative, we would specify values of *λ*_*s*_ that are at the *high end* of the range of plausible values. (b) These bounds should not be too far off because we expect the fraction *N*_*a*•_*/N*_*•*•_ to be fairly close to 1.

### Computation of sample size needed to test among asymptomatics

Specify the parameters listed in the section on **Prevalence testing** and then compute: Compute

- 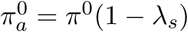 and similarly for 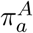
- 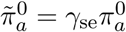and similarly for 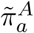
- Use binomial distribution on random variable 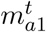 given *m*_*a*•_ to compute sample size *m*_*a*•_ needed to reject null hypothesis that 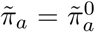 in favor of alternative that 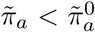 with 80% power given alternative 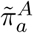.

### Symptomatic only testing

We can use a method similar to the foregoing to exploit *testing only in the population with COVID-like symptoms* to conduct rule-out hypothesis tests for prevalence above a maximally tolerable level *π*^0^. Assume that the proportion of the population with COVID-like symptoms is less than some value *ν*_*s*_, that is

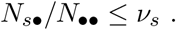

Note that with asymptomatic testing, we just operated on the bound that *N*_*a*•_ */N*_••_ ≤ 1. A comparable bound for *N*_*s*•_ */N*_••_ is not very sharp for symptomatic testing, hence the introduction of *ν*_*s*_. As above, this permits an upper bound on *π* as a function of *π*_*s*_:

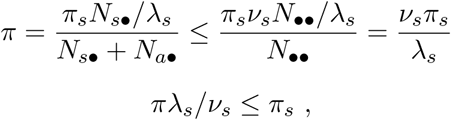

and we use this in the same way as we use *π*_*a*_. Namely, for a given null value *π*^0^ of *π*, we (conservatively) test whether *π*_*s*_ is less than a null value of *π*^0^*λ*_*s*_*/ν*_*s*_. If we reject, then, because

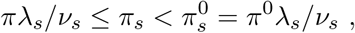

we will also reject *π* = *π*^0^ and conclude that *π < π*^0^. Remarks: (a) Here, we would select *λ*_*s*_ at the *low end* of the range of plausible values, to be conservative. (b) Accordingly, we also select *ν*_*s*_ at the high end of plausible values.

### Computation of sample size needed to test among symptomatics

Specify the parameters in the section on **Prevalence testing** and then compute:

- 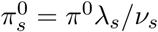and similarly for 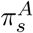.
- 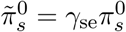and similarly for 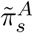
- Use binomial distribution on random variable 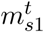 similarly to the plan for the asymptomatic testing.

## Data Availability

There are no data in this manuscript. We are happy to provide R code for generating the results.

## Acknowledgement

The authors thank Elizabeth C. Matsui in the Department of Population Health at University of Texas at Austin for exchanges helping to frame the problem.

## Footnotes

1 In statistical terms, one minus the Type I error rate in a one-sided hypothesis test of *π* = *π*^0^ versus *π < π*^0^.

2 In statistical terms, this is the power of the hypothesis test.

3 Actual power is greater than the target owing to the discreteness of the problem.

## References

Poletti et al. (2020). Probability of symptoms and critical disease after SARS-CoV-2 infection. arXiv:2006.08471v2.

Eggo et al. Respiratory virus transmission dynamics determine timing of asthma exacerbation peaks: Evidence from a population-level model. PNAS February 23, 2016 113 (8) 2194–2199.

Centers for Disease Control and Prevention. Estimated Influenza Illnesses, Medical visits, Hospitalizations, and Deaths in the United States — 2018–2019 influenza season. January 8, 2020.

Martin et al. Modelling testing frequencies required for early detection of a SARS-CoV-2 outbreak on a university campus. medRxiv 2020: https://doi.org/10.1101/2020.06.01.20118885.

Gostic et al. Estimated effectiveness of symptom and risk screening to prevent the spread of COVID-19. eLife 2020;9.

